# Basivertebral nerve block during vertebral augmentation: An alternative approach to intraprocedural pain management

**DOI:** 10.1101/2021.02.27.21251560

**Authors:** Giovanni C. Santoro, Siddhant Kulkarni, Kenny Lien

**Author notes:** **Corresponding Author:** Giovanni C. Santoro DO, Interventional Radiology Resident Physician; Northwell Health – Mather Hospital, 75 N Country Road, Port Jefferson, NY 11777.

## Abstract

Vertebral compression fractures secondary to osteoporosis can be treated with vertebral augmentation. Since intraprocedural pain is common during vertebral body endplate manipulation, these procedures are often carried out using conscious sedation or general anesthesia. Research has shown that the vertebral endplates are innervated by the basivertebral nerve, which has been successfully targeted via radiofrequency ablation to treat chronic vertebrogenic lower back pain. With this physiology in mind, we treated ten patients with vertebral compression using intraosseous basivertebral nerve block as the primary intraprocedural analgesia. In this case series, we describe our successful experience with this novel approach.

## Introduction

Vertebral compression fractures (VCF) of the thoracic and lumbar spine secondary to osteoporosis are often treated with percutaneous vertebral augmentation (PVA). Using fluoroscopic guidance, percutaneous access into the selected vertebral level is obtained via transpedicular or extrapedicular advancement of an introducer needle. Once within the vertebral body, a cavity is created with a curette device and often with balloon augmentation. Cement is then administered under fluoroscopic guidance through the introducer needle into the cavity. During the procedure, the patient may experience significant pain due to vertebral body morphologic change and radiating pressure on the cortical bone of the superior and inferior endplates. Therefore, PVA is performed under conscious sedation in most cases. However, at certain institutions, general anesthesia is often used.

The overall risk for single-level treatment remains low to moderate [1, 2]. However, there exists a subset of patients that would benefit clinically from augmentation, but are considered high risk candidates for anesthesia or sedation due to medical comorbidities. Contraindications to anesthesia, may result in certain VCFs being left untreated, which may have detrimental enduring effects. Long term studies have demonstrated that patients with VCF who do not receive PVA, have decreased mobility, increased morbidity and mortality, and an overall decreased quality of life [2-5]. Therefore, if possible, it would be clinically valuable to reduce the procedural risk in this population, and offer vertebral augmentation to these patients.

The basivertebral nerve (BVN) innervates the superior and inferior endplate of the vertebral bodies [6-8]. Recent histologic and clinical evidence suggests that axial load pain directed at the vertebral endplates is significantly reduced by ablation of this nerve [9]. Permanent ablation of the BVN has proven successful in treating chronic vertebrogenic lower back pain via the Intracept® procedure [9, 10]. Leveraging this knowledge, we postulated that temporary intraosseous BVN block could be used as an alternate intraprocedural analgesia during PVA, thereby eliminating the need for conscious sedation or general anesthesia.

## Material & Methods

This study was performed at a single center community-based academic hospital interventional radiology practice, where vertebral augmentation is performed on a regular basis by a provider with experience performing the Intracept® procedure. A request for human subjects research approval was submitted to the local Institutional Research Board (IRB) at Mather Hospital and to the Human Research Protection Program at the Feinstein Institute for Medical Research at our parent organization of Northwell Health. Approval was obtained and research was carried out in accordance with the IRB. A total of ten patients (five female, five male) between ages 50-90 years old were consented and enrolled in this study. All ten patients were of Caucasian ethnicity. Our inclusion criteria required that patients had a diagnosis of osteoporosis on dual energy x-ray absorptiometry (DEXA), had either an acute or subacute single level VCF between T10 – L3 as confirmed via magnetic resonance imaging (MRI) or nuclear medicine bone scan, and had initial pain score of greater than or equal to five upon initial consultation. The pain score used was the standard clinical Visual Analog Scale (VAS) of 0 – 10, where zero indicates no pain and ten indicates the worst pain of the patient’s life. Patients with psychiatric comorbidities, including anxiety, depression, and psychosis, were excluded from the study to mitigate the interaction of long-term sedative and anxiolytic medication use. Patients with a history of illicit drug abuse and/or alcohol abuse/dependence were excluded to minimize possible confounding effects due to altered nociception among these patients. Patient with Parkinson’s Disease or other movement disorders were also excluded to minimize any potential confounding effects of dopaminergic medications.

All procedures were carried out by a single attending interventional radiologist. All patients received routine periprocedural clinical care, as well as additional intraprocedural monitoring for pain. During each procedure, local anesthesia was achieved using subcutaneous and periosteal injection of 2% lidocaine solution. Thereafter, the affected vertebral body was accessed by advancing an 11-guage trocar introducer needle via a posterior transpedicular approach under continuous fluoroscopic guidance. The introducer needle was positioned just anterior to the posterior wall of the vertebral body, as confirmed on lateral imaging. The needle was then replaced with a curved cannula and was intermittently advanced under fluoroscopic guidance using alternating anteroposterior (AP) and lateral views, until the curved cannula was positioned at the expected anatomic location of the BVN. The positioning of the cannula was determined to be adequate once the distal end was centrally located within the vertebral body on AP imaging and between 30-50% of the anterior-to-posterior length of the vertebral body from the posterior wall on lateral view (Figure 1). Once positioning was confirmed, an intraosseous injection of 5 ml of 2% lidocaine solution was instilled via the introducer to establish nerve block. Thereafter, vertebral augmentation was carried out using a unipedicular approach, as per operator routine.

**Figure 1.**
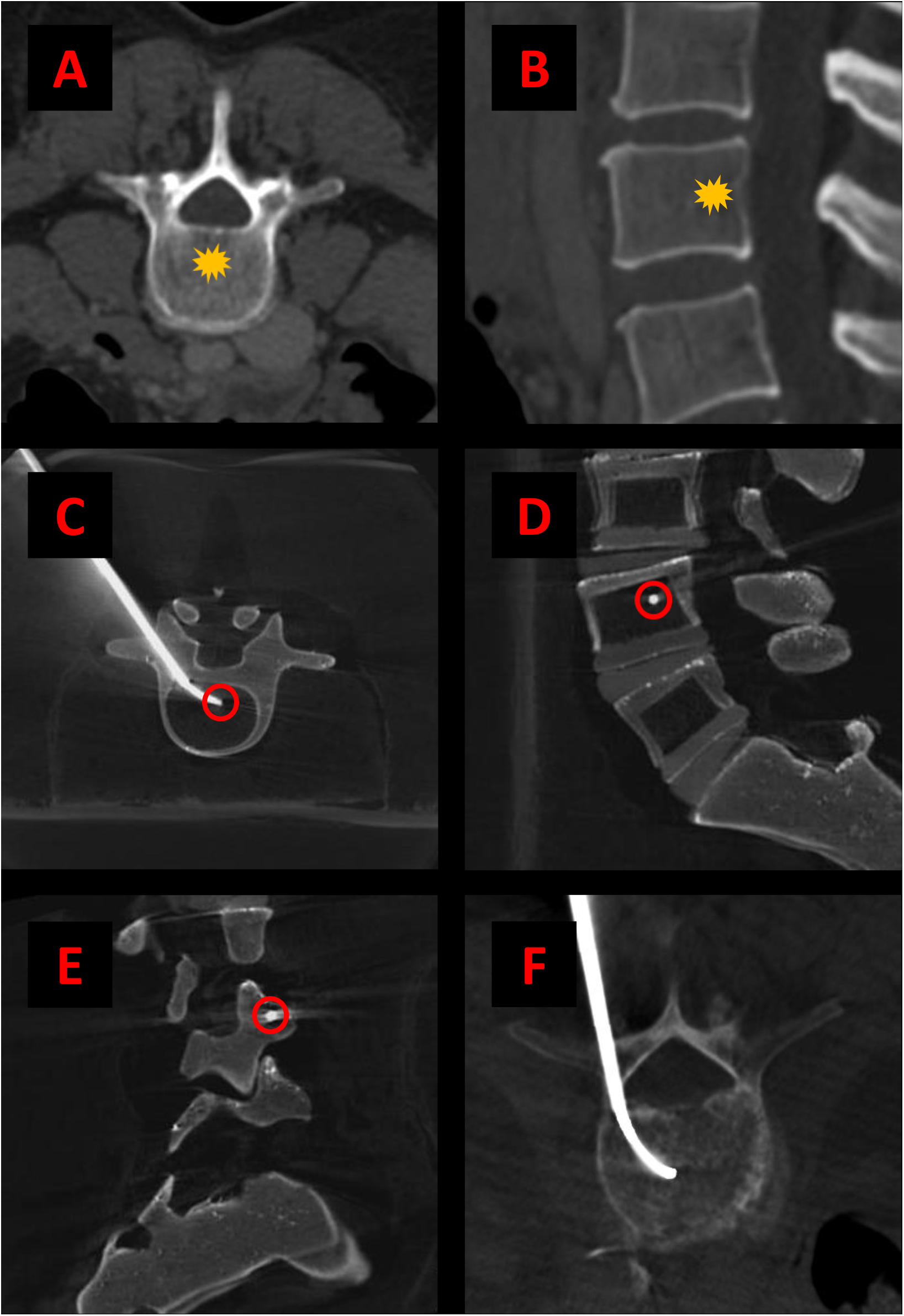
Example axial (A) and sagittal (B) CT images demonstrating the expected location of the basivertebral nerve (yellow star). Example cone-beam CT images acquired using a medtronic kyphone® training model showing transpedicular curved canula placement on axial (C), lateral (D), and oblique lateral (E) Views. Canula tip placement (red circles) is ideally centrally located within the vertebral body on AP imaging and between 30-50% of the anterior-to-posterior length of the vertebral body from the posterior wall on lateral view. Example showing curved canula placement in a patient undergoing kyphoplasty with BVN block (F).

In addition to intermittent intraprocedural pain monitoring, patient pain was assessed using the VAS in the preoperative period, in the immediate postoperative period (following transfer of the patient from the fluoroscopy suite to the transport stretcher) and during routine 1 week follow up in clinic. Patients were also asked to rate their satisfaction using a subjective Likert scale of 1 – 4, at the same time intervals. Satisfaction ratings were defined as follows: 1 = completely dissatisfied, 2 = dissatisfied, 3 = satisfied, 4 = very satisfied. Upon discharge, patients were not prescribed opioid pain relievers by the interventional radiology medical staff. Patients were counseled to use over-the-counter analgesics, such as acetaminophen or nonsteroidal anti-inflammatory drugs, as needed.

## Results

All ten patients successfully underwent vertebral augmentation using basivertebral nerve block as the primary intraoperative anesthesia. None of the patients enrolled in the study required additional analgesia or subsequent intraprocedural sedation. Routine monitoring of vitals did not show any significant fluctuation in blood pressure or heart rate, as defined as greater than 20% deviation from baseline.

Pain scores, satisfaction scores, and additional data are reported in Table 1. Nine of ten patients reported a complete reduction in pain immediately after their procedure. Only one patient reported 3/10 pain immediately post-procedure, which was overall reduced from the patient’s baseline of 8/10. This patient subsequently demonstrated a further reduction in pain at one week, reporting 2/10 pain at that time. At one-week follow-up, one patient reported 3/10 pain and three patients reported 2/10 pain, while the remaining six patients reported no pain. Notably, all patients overall reported a decrease in pain score at one-week follow-up compared to their baseline at initial consultation. All patients reported a 4/4 (very satisfied) satisfaction score both immediately after their procedure and at one-week follow-up.

**Table 1.** Summary table includes relevant patient demographics. Pain scores (PS) were obtained during preoperative consultation (pre), during the immediate postoperative period (post), and during the patient’s one-week clinical follow-up (1 wk.). Satisfaction scores (SS) were also obtained postoperatively (post) and at one-week follow-up (1 wk.). Routine intraprocedural monitoring of vitals was carried out. There were no disturbances in patient blood pressure or heart rate as defined as >20% deviation from baseline (BP/HR Δ). Intraprocedural pain monitoring was routinely carried out, with special attention to intraprocedural balloon augmentation (Balloon Pain). No significant pain was reported by any patient during balloon inflation. Intravertebral cement injection was carefully carried out under fluoroscopic guidance to monitor for potential extravasation. The total volume of cement administered to each patient was recorded (Cement Vol.)

| Patient | Sex | Fx Level | PS (pre) | PS (post) | PS (1 wk.) | SS (post) | SS (1 wk.) | BP/HR Δ | Balloon Pain | Cement Vol. (mL) |
| --- | --- | --- | --- | --- | --- | --- | --- | --- | --- | --- |
| 1 | FM | L1 | 8 | 0 | 0 | 4 | 4 | none | none | 3.5 |
| 2 | FM | T11 | 10 | 0 | 0 | 4 | 4 | none | none | 1.5 |
| 3 | FM | T10 | 8 | 0 | 3 | 4 | 4 | none | none | 3.5 |
| 4 | FM | T11 | 9 | 0 | 0 | 4 | 4 | none | none | 4.0 |
| 5 | FM | L2 | 8 | 3 | 2 | 4 | 4 | none | none | 3.0 |
| 6 | M | L3 | 5 | 0 | 0 | 4 | 4 | none | none | 3.5 |
| 7 | M | L1 | 8 | 0 | 2 | 4 | 4 | none | none | 3.0 |
| 8 | M | L1 | 6 | 0 | 0 | 4 | 4 | none | none | 3.5 |
| 9 | M | L5 | 9 | 0 | 0 | 4 | 4 | none | none | 3.5 |
| 10 | M | L2 | 5 | 0 | 2 | 4 | 4 | none | none | 3.5 |

## Discussion

The goal of our study was to determine the feasibility of performing vertebral augmentation without conscious sedation or general anesthesia, and instead by using only local anesthesia and basivertebral nerve block. This case series suggests that this is both safe and effective. The results of the study are consistent with research demonstrating the efficacy of basivertebral nerve ablation for chronic vertebrogenic lower back pain.

Reviews comparing non-operative and operative management of vertebral compression fractures, demonstrate the efficacy of vertebral augmentation in reducing morbidity and mortality risk. Percutaneous vertebral augmentation is a minimally invasive procedure with relatively low rates of major complications. It is widely offered in many outpatient and inpatient settings. However, there is a subset of patients with pathologic vertebral compression fractures and comorbidities such as heart failure or severe emphysema, who do not undergo the procedure due to the increased risk associated with sedation or general anesthesia. We believe that by providing alternative intraoperative analgesia in these high-risk patients, we are better able to serve this population.

Although our study is limited due to the lack of control groups and long-term follow-up, we were able to successfully eliminate the need for general anesthesia or conscious sedation in these select patients. This alone demonstrates that we can reduce the procedural risk associated with anesthesia in vertebral compression fracture patients, who are generally older and who often have multiple medical comorbidities. To our knowledge, this is the first report where basivertebral nerve block has been used during vertebral augmentation. We hope that this positive experience will foster future research and reduce procedural risk, thereby increasing the quality of life among select patients with vertebral compression fractures.

## Data Availability

All data is available upon request and at the discretion of the investigators and physicians.

